# Development of a Nomogram for Predicting the Risk of Hospital-Acquired Pressure Injuries in Patients in the Cardiovascular Intensive Care Unit

**DOI:** 10.1101/2023.11.14.23298510

**Authors:** Hao-yue Li, Yan Zhang, Xiao-lan Shui, Zhi-gang Che, Qi-fan Liu, Shan-shan Guo

**Author notes:** Correspondence Shan-shan Guo, Renmin Hospital of Wuhan University, 238 Jiefang Road, Wuhan, 430060, China. H.L. and Y.Z. are co-first author. Conflicts of Interest. The authors have no conflicts of interest to disclose.

## Abstract

**Purpose:** Hospital-acquired pressure injuries (HAPIs) increase the medical burden of patients in the cardiovascular intensive care unit (CCU). Thus, identification of CCU patients with a risk for HAPIs is important. To establish a nomogram model for predicting the occurrence of HAPIs in patients in the CCU.

**Methods:** This was a retrospective cohort study in patients in the CCU at our hospital who developed HAPIs between January 2023 and June 2023. Patient data were extracted from the hospital’s information management system. Risk factors for HAPIs were identified using univariate and multivariate logistic regression analyses and the screened indicators were integrated into a nomogram. The effectiveness of the nomogram was evaluated and verified using receiver operating characteristic curve and decision curve analysis (DCA).

**Results:** In this study, a total of 161 patients were enrolled. Univariate logistic regression analysis showed that vasopressor use (OR=2.51, 95%CI=1.24–5.09, *p*=.010) and NT-proBNP (OR=2.92, 95%CI=1.45–5.92, *p*=.003), lactic acid (OR=9.43, 95%CI=4.15–21.41, *p* <.000), procalcitonin (OR=1.37, 95%CI=1.07–1.75, *p*=.012), D-dimer (OR=1.16, 95%CI=1.05–1.28, *p*= .004), and albumin (OR=0.88, 95%CI=0.81–0.95, *p*=.002) levels were independent risk factors for HAPIs. Multivariate logistic regression analysis showed that vasopressor use (OR=3.05, 95%CI=1.20–7.73, *p*=.019) and lactic acid (OR=12.05, 95%Cl=4.13–35.21, *p* <.000), procalcitonin (OR=1.30, 95%Cl=1.01–1.69, *p*=.043) and albumin (OR=0.82, 95%Cl=0.73–0.93, *p*=.002) levels were independent risk factors for HAPIs. The nomogram was well-calibrated and showed good discriminative ability (AUC=.87). The DCA showed a better net benefit, and the results were confirmed within the validation cohort.

**Conclusion:** The nomogram model developed in this study showed good predictability and can identify patients at risk of developing HAPIs and aid the formulation of targeted interventions.

## INTRODUCTION

In 2016,the National Pressure Ulcer Advisory Panel has defined a pressure injury as localized damage to the skin or underlying soft tissues. Pressure ulcers that develop after a patient has been hospitalized are referred to as hospital-acquired pressure injuries (HAPIs), and they are typically the result of pressure and/or shear forces.^1^ Tissue microenvironment, complications, perfusion, and soft tissue conditions affect the tolerance of local tissues to pressure and shear stress.^2^ The incidence of pressure ulcers in hospitalized patients is mainly concentrated in the intensive care unit (ICU) and in the emergency, geriatric, orthopedic, neurology, and rehabilitation departments, with the emergency and critical care departments having the highest incidence.

The incidence of pressure ulcers in the ICU ranges from 1% to 56%, which is 2-3 times that in the general ward.^3^ This high incidence of pressure ulcers in ICUs is attributable to multiple reasons, such as severe, complex, and changeable conditions; hemodynamic instability; internal environment disorder; and mechanical assistance.^4^ The cardiovascular care unit (CCU), a specialized ICU, covers the treatment of a variety of critical cardiovascular diseases, such as cardiogenic shock, acute myocardial infarction, viral myocarditis, malignant arrhythmia, and acute heart failure. The CCU is characterized by the management of critical and complex conditions and rapid changes. Patients in the CCU are exposed to multiple risk factors, and the occurrence of pressure ulcers can increase their medical burden. The initial step in the management of pressure ulcers is conducting a risk assessment; therefore, accurate identification of risk factors and high-risk patients using standard and effective risk assessment tools is essential.^5^ In clinical practice, various pressure ulcer risk assessment scales are frequently employed, including the Braden, Norton, and Waterlow scales.^6–9^ The high sensitivities of these scales have been verified; however, some studies revealed that they show high false-positive rate and low specificity for ICU patients. The Conscious level, Mobility, Hemodynamics, Oxygenation, Nutrition index is commonly used for pressure ulcer risk assessment in the ICU owing to its predictive ability and relatively fixed critical values.

The evaluation items of pressure ulcer risk assessment scales rely on the judgment of clinical nurses and are based on dynamic behaviors and the physical function statuses of patients. These subjective observations and evaluations are easily affected by individual differences. In addition, these assessment tools are universal scales, and their diagnostic effects focus on a single disease rather than on a system, such as the cardiovascular system. These factors limit the use of these scales for pressure ulcer management in CCU patients. The primary risk factors for pressure ulcers among patients in different ICUs, such as the RICU, UICU, CCU, and NICU, differ because of the different characteristics of the critical diseases treated in these units.^10^ Recent studies have demonstrated that clinical indicators, such as hemodynamics, internal environment, and use of some drugs, are closely related to skin status changes. We believe that a combination of these objective clinical indicators may be more effective than subjective evaluation in predicting the risk of pressure ulcers among patients in cardiovascular departments. Therefore, we conducted research to develop a pressure ulcer risk prediction model suitable for CCU patients. This prediction model is a dynamic nomogram with visibility and mathematical advantages that can integrate multiple prognostic and determinant variables to aid clinical decision-making and produce numerical probabilities of clinical events that can be used for patient evaluation. More importantly, we validated that the nomogram is beneficial for timely identification, prevention, and treatment of pressure injuries in CCU patients. Accurate evaluation, timely prevention, and appropriate intervention measures are essential for reducing pressure ulcer incidence in the CCU.

## METHODS

### PARTICIPANTS

A retrospective analysis was conducted on the clinical data of patients admitted to the CCU of Renmin Hospital of Wuhan University from January 2023 to June 2023. The inclusion criteria were as follows: (1) no pressure sores on the skin on CCU admission; (2) age >18 years; and (3) continuous CCU stay for >24 hours. The exclusion criteria were as follows: (1) patients transferred from other departments; (2) patients with malignant cachexia; (3) patients who had pressure ulcers before they were admitted to the hospital; and (4) patients with severe skin diseases.

### DATA COLLECTION

All data in this study were extracted from the available information management system of our hospital. The data extracted included age; sex; history of diabetes; ejection fraction (EF) on echocardiography; blood pressure; creatinine; albumin (Alb); brain natriuretic peptide (BNP); procalcitonin (PCT); hemoglobin (HB); lactic acid (Lac); and D-dimer levels; the utilization of mechanical assist devices, such as extracorporeal membrane oxygenation (ECMO), intra-aortic balloon pumps, and ventilators; vasopressor use; and diuretic use. Information on the skin condition of patients with pressure injuries was extracted using the nursing management system of our hospital. The management of pressure injury in our hospital involves completing skin examination and assessment of patients when they are admitted to the hospital, and conducting a Braden scale assessment within 4 h. Patients with a high risk of developing pressure ulcers are predicted in the nursing management system, and preventive measures, such as turning over every two hours, enhancing nutrition, and keeping the skin clean and dry, are initiated. In our hospital, skin identification is performed by wound stoma nursing experts. Pressure ulcer records are filled in the nursing management system, and dynamic tracking management is performed.

### CONSTRUCTION OF THE NOMOGRAM MODEL

Variables with *p* <.05 in the univariate regression analysis were included in the multivariate regression analysis. Each factor was estimated separately, and a nomogram for assessing the risk for HAPIs was constructed. A receiver operating characteristic (ROC) curve was employed to assess the nomogram’s predictive accuracy for the risk of HAPIs. Calibration curves were used to depict the predicted and observed probabilities of the nomogram. Decision curve analysis (DCA) demonstrated potential net benefits. The discriminative performances of EF and HB, PCT, and Alb levels were compared with that of the nomogram.

### STATISTICAL ANALYSIS

Quantitative variables were expressed as either mean ± standard deviation or median (interquartile range), and comparisons were made using an unpaired two-tailed Student’s t-test or Kruskal-Wallis test. Categorical data were presented as numbers (percentages) and were compared using the chi-square test or Fisher’s exact test, as applicable. Univariate and multivariate logistic regression analyses were conducted using SPSS software, version 26. Nomograms were constructed using the "rms" package in the R software. ROC and DCA analyses were performed using the "pROC" and "rmda" packages.

#### Ethical considerations

This study received approval from the Ethics Committee of Renmin Hospital of Wuhan University. As the study was designed as a retrospective review of the data, the requirement for informed consent was waived.

## RESULTS

A total of 161 CCU patients were enrolled in this study between January 1, 2023, and June 1, 2023. There were 52 patients with HAPIs. The participants were divided into a training group (n = 109) and a validation group (n=52). The clinical and laboratory characteristics of the patients are shown in table 1. Regarding HAPIs, 61 of the 161 patients (37.9%) had HAPIs. Of these, 41 (37.6%) and 20 (38.5%) were in the training and validation groups, respectively. In the training set, univariate logistic regression analysis revealed that vasopressor use (OR= 2.51, 95%CI=1.24–5.08, *p*=.010) and N-terminal pro–B-type natriuretic peptide (NT-proBNP) (OR=2.93, 95%CI=1.45–5.91, *p*=.003), Lac (OR=9.43, 95%CI=4.15–21.41, *p*< .001), PCT (OR= 1.37, 95%CI=1.07–1.75, *p*=.012), D-dimer (OR=1.16, 95%CI=1.05–1.28, *p*= .004), and Alb (OR=0.88, 95%CI=0.81–0.95, *p*=.002) levels were independent risk factors for HAPIs. Multivariate logistic regression analysis revealed that vasopressor use (OR=3.05, 95%CI=1.20–7.73, *p*=.019) and Lac (OR=12.05, 95%Cl=4.13–35.21, *p*< .001), PCT (OR=1.30, 95%Cl=1.01–1.69, *p*=.043), and Alb (OR=0.82, 95%Cl=0.73–0.93, *p*=.002) levels were independent risk factors for HAPIs(table 2).

**Table 1.** Baseline Characteristics of the Patients.

| Variable | Training cohort | Validation cohort | *P* value |
| --- | --- | --- | --- |
| Age (n [%]) |  |  | .238 |
| < 70 | 35 | 12 |  |
| ≥70 | 74 | 40 |  |
| Gender (n [%]) |  |  | .605 |
| Men | 66 | 34 |  |
| Women | 43 | 18 |  |
| Hypotension (n [%]) |  |  | .864 |
| Yes | 44 | 20 |  |
| No | 65 | 32 |  |
| EF (n [%]) |  |  | .731 |
| < 40% | 68 | 34 |  |
| ≥40% | 41 | 18 |  |
| Hb(n [%]) |  |  | .279 |
| < 110g/L | 37 | 13 |  |
| ≥110g/L | 72 | 39 |  |
| NT-pro BNP(n [%]) |  |  | .675 |
| < 5000pg/mL | 59 | 28 |  |
| ≥5000 pg/mL | 50 | 24 |  |
| Cr (n [%]) |  |  | .129 |
| < 111μmol/L | 57 | 20 |  |
| ≥111μmol/L | 52 | 32 |  |
| Lac(n [%]) |  |  | .406 |
| < 1.5mmol/L | 56 | 23 |  |
| ≥1.5 mmol/L | 53 | 29 |  |
| PCT [M (P25 ,P75 ) , ng/mL ] | 0.25(0.09,1.12) | 0.20(0.09,0.65) | .663 |
| D-dimer[M (P25 ,P75 ) , mg/L] | 1.61(0.70,3.30) | 1.73(0.86,2.85) | .697 |
| Alb (g/L) | 37.01±7.04 | 38.47±9.62 | .327 |
| Use of diuretics (n [%]) |  |  | >.999 |
| No | 29 | 10 |  |
| Yes | 80 | 42 |  |
| Mechanical assistance (n [%]) |  |  | 0.517 |
| No | 74 | 36 |  |
| Yes | 35 | 16 |  |
| Diabetes (n [%]) |  |  | 0.858 |
| No | 35 | 18 |  |
| Yes | 74 | 34 |  |
| Use of vasopressors |  |  | 0.868 |
| No | 52 | 24 |  |
| Yes | 57 | 28 |  |

EF, left ventricular ejection fraction; Hb, hemoglobin; Cr, creatinine; Lac, lactic acid; PCT, procalcitonin

**Table 2.**
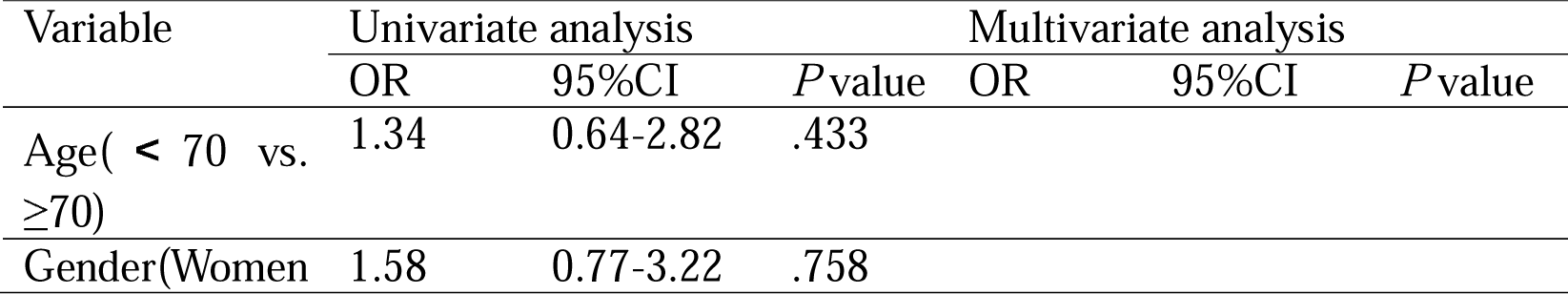
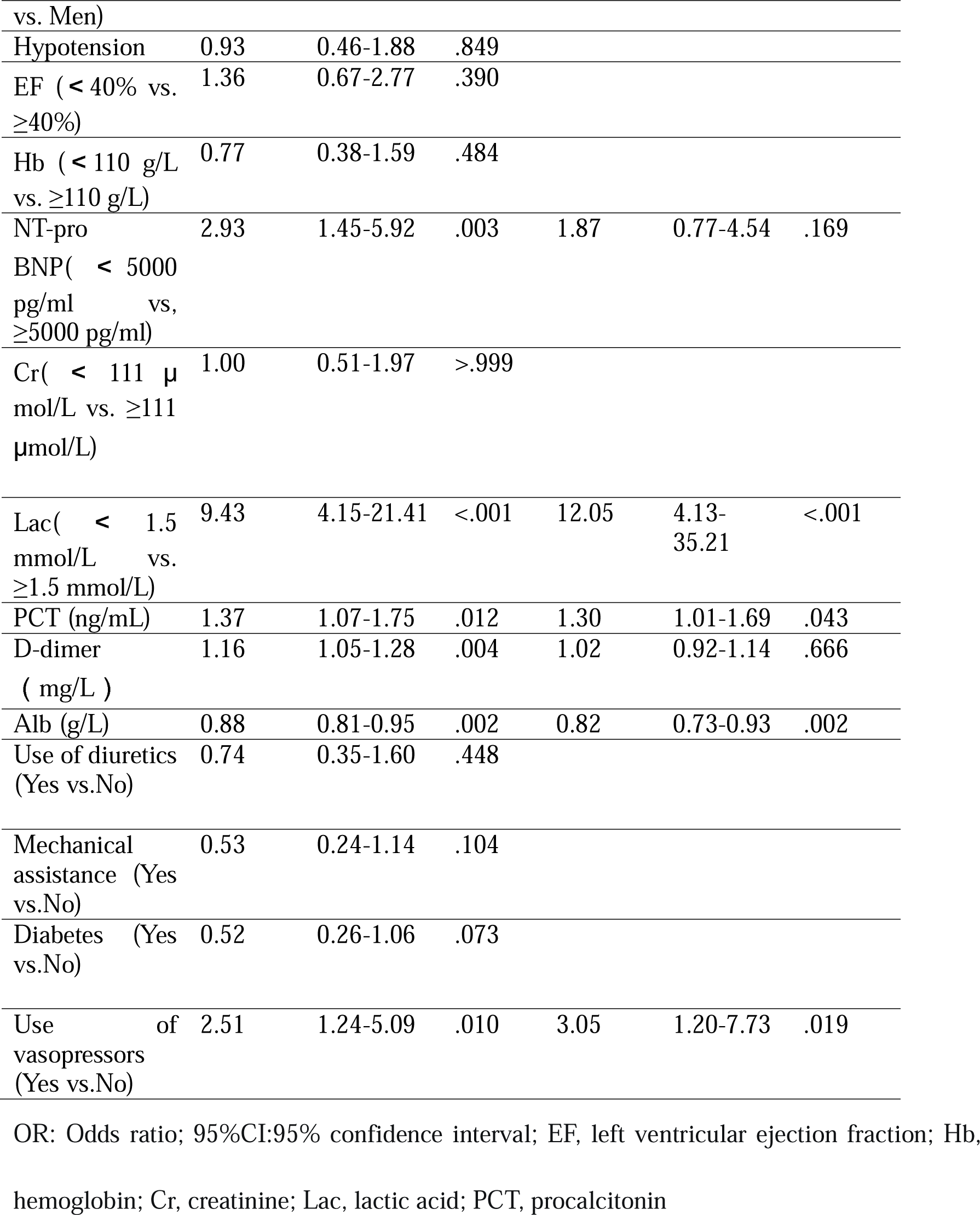
Univariate and Multivariate Logistic Regression Analyses of the Risk of HAPIs in CCU Patients.

| Variable | Univariate analysis |  |  | Multivariate analysis |  |  |
| --- | --- | --- | --- | --- | --- | --- |
|  | OR | 95%CI | *P* value | OR | 95%CI | *P* value |
| Age( < 70 vs. ≥70) | 1.34 | 0.64-2.82 | .433 |  |  |  |
| Gender(Women | 1.58 | 0.77-3.22 | .758 |  |  |  |
| vs. Men) |  |  |  |  |  |  |
| Hypotension | 0.93 | 0.46-1.88 | .849 |  |  |  |
| EF ( < 40% vs. ≥40%) | 1.36 | 0.67-2.77 | .390 |  |  |  |
| Hb ( < 110 g/L vs. ≥110 g/L) | 0.77 | 0.38-1.59 | .484 |  |  |  |
| NT-pro BNP( < 5000 pg/ml vs, ≥5000 pg/ml) | 2.93 | 1.45-5.92 | .003 | 1.87 | 0.77-4.54 | .169 |
| Cr( < 111 μmol/L vs. ≥111 μmol/L) | 1.00 | 0.51-1.97 | >.999 |  |  |  |
| Lac( < 1.5 mmol/L vs. ≥1.5 mmol/L) | 9.43 | 4.15-21.41 | <.001 | 12.05 | 4.13-35.21 | <.001 |
| PCT (ng/mL) | 1.37 | 1.07-1.75 | .012 | 1.30 | 1.01-1.69 | .043 |
| D-dimer ( mg/L ) | 1.16 | 1.05-1.28 | .004 | 1.02 | 0.92-1.14 | .666 |
| Alb (g/L) | 0.88 | 0.81-0.95 | .002 | 0.82 | 0.73-0.93 | .002 |
| Use of diuretics (Yes vs.No) | 0.74 | 0.35-1.60 | .448 |  |  |  |
| Mechanical assistance (Yes vs.No) | 0.53 | 0.24-1.14 | .104 |  |  |  |
| Diabetes (Yes vs.No) | 0.52 | 0.26-1.06 | .073 |  |  |  |
| Use of vasopressors (Yes vs.No) | 2.51 | 1.24-5.09 | .010 | 3.05 | 1.20-7.73 | .019 |

OR: Odds ratio; 95%CI:95% confidence interval; EF, left ventricular ejection fraction; Hb, hemoglobin; Cr, creatinine; Lac, lactic acid; PCT, procalcitonin

### PERFORMANCE OF THE NOMOGRAM

Variables with a *p* value <0.05 in both the univariate and multivariate logistic regression analyses were included in a nomogram for predicting the risk for HAPIs ((figure 1). The nomogram was then used to predict the occurrence of HAPIs (Lac level, 1.70 mmol/L; Alb level, 32.50 g/L; PCT level, 3.20 ng/mL; vasopressor use). The score for Lac level, Alb level, PCT level, and vasopressor use in the nomogram was 20, 80, 5, and 8, respectively. The total score of the HAPI risk prediction nomogram, which is the sum of all the above-mentioned scores, was 113. Ultimately, the risk of HAPI was estimated to exceed 90.0%.

**Figure 1.**
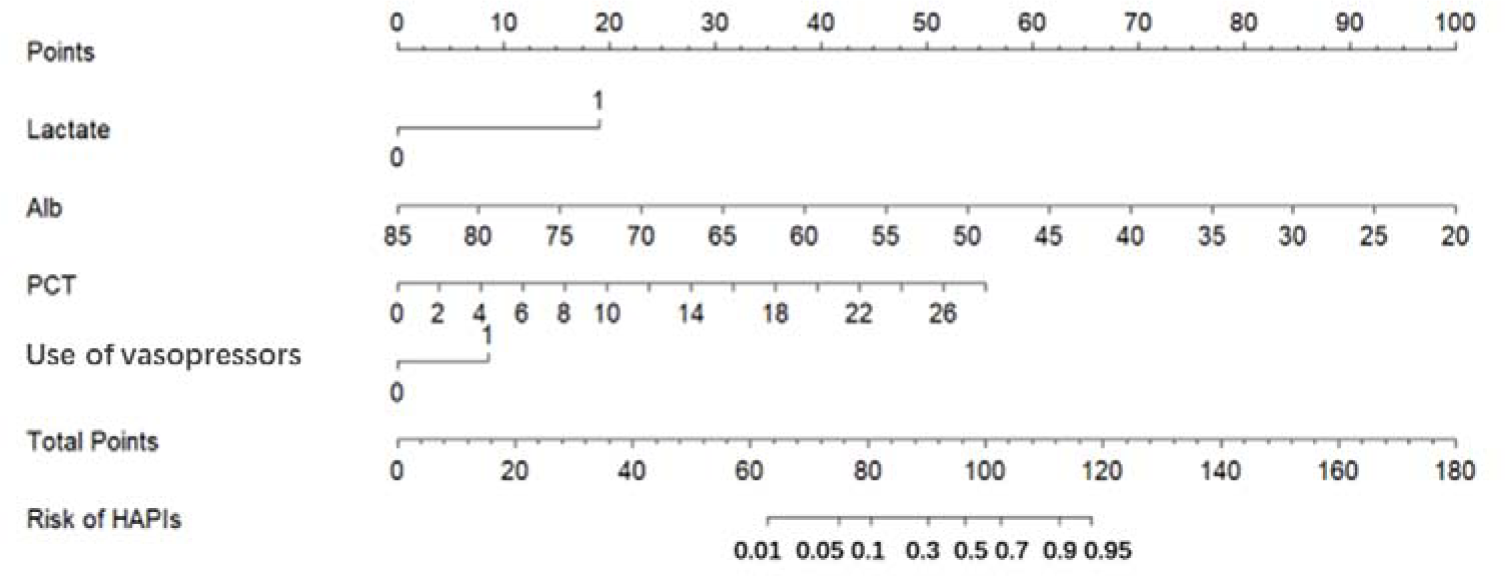
Nomogram for predicting the risk of HAPI. PCT procalcitonin; Alb, albumin

### COMPARISON OF THE NOMOGRAM AND OTHER INDICES

As shown in figure 2, the calibration curves of the nomogram closely matched the standard curves of the training and validation cohorts, signifying that the plot was well-calibrated. The AUCs of the HAPI risk prediction nomogram in the training and validation groups were 0.868 and 0.852, respectively. These values were superior to those of other indices. DCA is a new prediction tool used to evaluate the validity of nomograms. The decision curves for the nomogram and other indices for the two cohorts can be seen in Figure 3. These results suggest that the nomogram offered more reliable clinical guidance compared to the other indices.

**Figure 2.**
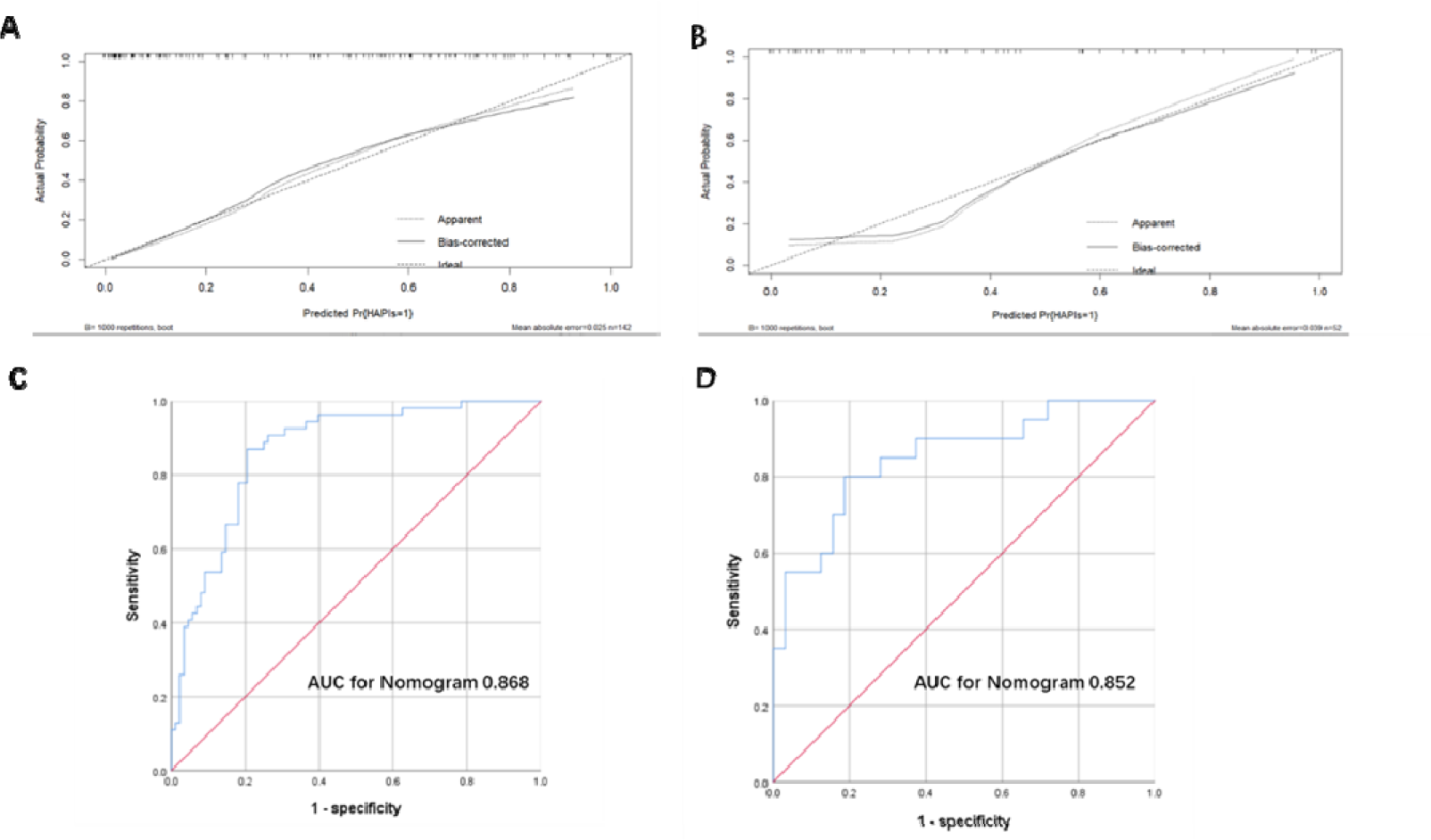
Calibration plots and the areas under the curves (A) The calibration curve of the nomogram in the training cohort. (B) The calibration curve of the nomogram in the validation cohort. (C) The areas under the curves (AUCs) of the HAPI risk prediction nomogram in the training cohort. (D) The areas under the curves (AUCs) of the HAPI risk prediction nomogram in the validation cohort.

**Figure 3.**
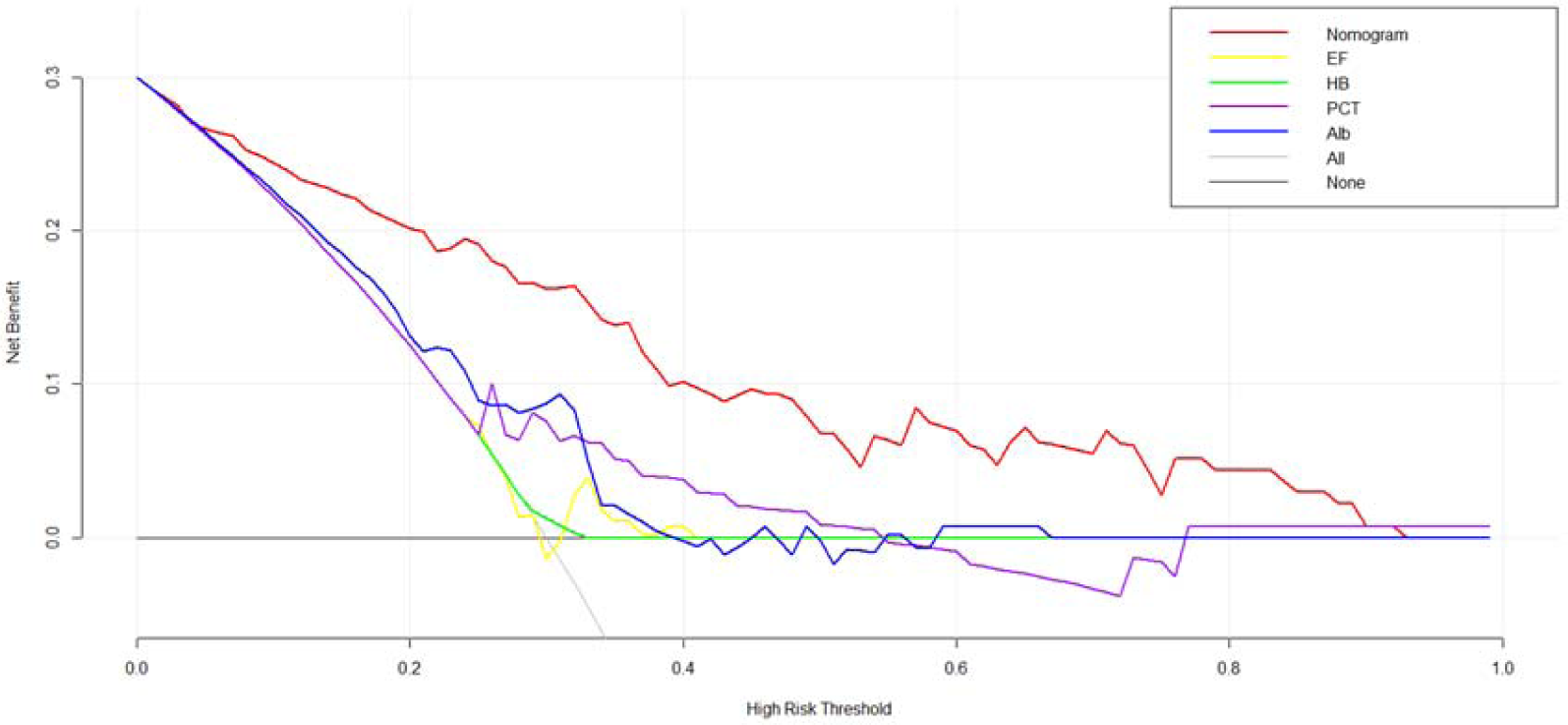
Decision curves of the nomogram in the training cohort. EF, ejection fraction; HB, hemoglobin; PCT procalcitonin; Alb, albumin

## DISCUSSION

According to a survey conducted in Germany, the incidence of pressure ulcers ranges from 5.0% to 12.5%,^11^ with incidences of 8.9%, 22.9%, and 18.5% reported in France, Canada, and Ireland, respectively. Groeneveld et al. reported that the prevalence of pressure ulcers in the ICU is 26.3%.^12^ The incidence of pressure ulcers in China is much lower than those reported in foreign countries. The reason for this may be differences in pressure ulcer reporting mechanisms and omission and concealment of some cases.^13^ Existing research on the occurrence of pressure ulcers in hospitals are mostly limited to evaluation of the declaration system and the formulation of management processes, most of which are focused on the introduction of management experience and lack of clinical epidemiological research on risk factors for pressure ulcers. Moreover, assessment of pressure ulcer risk tools used in various hospitals differ, and the evaluation of risk grade is inconsistent and lacks accuracy and objectivity, which can easily cause excessive or insufficient nursing.

The CCU is different from the ICU. Although patients in the CCU are bedridden, most of them are conscious, can eat independently, and can turn over and change their positions in bed. The present study showed that vasopressor use and Lac, Alb, and PCT levels were strongly associated with the occurrence of pressure ulcers in the CCU, and had good predictive value for HAPIs.

Nutrition is evaluated in some domains of pressure ulcer assessment scales. However, even with a normal diet intake, patients with different body types and low or high BMI may have nutrient deficiencies. Therefore, the nutritional statuses of patients cannot be assessed using BMI or dietary status alone. We believe that a combination of objective clinical test indicators can accurately reflect the nutritional status of patients. Nutritional laboratory markers include serum Alb, HB, prealbumin, zinc, and transferrin levels. Of these, serum Alb is the most accurate indicator of the long-term nutritional status of patients.^14^ Alb level is not only indicative of the nutritional status but also indicates a response to disease status. Heart failure patients admitted to the CCU often show hypoproteinemia, which is confirmed by measuring protein levels, including albumin levels. Hypoproteinemia affects cardiac function and prognosis in patients with heart failure. Alb levels are also strongly associated the occurrence of pressure ulcers. Low Alb levels can cause skin edema. Edema in the interstitial space can reduce capillary blood flow and affect the skin’s oxygen supply, which further threatens the skin’s nutrient supply and makes it more vulnerable to damage.^15^ Serra et al. found that hypoalbuminemia is significantly correlated with the occurrence of pressure ulcers^16^ when the total blood protein level is lower than 6 g/L. In this condition, treatment of pressure ulcers will not have a curative effect.^17^ Alb has a high molecular weight, which can effectively maintain the osmotic pressure of normal plasma colloid, thus regulating the water dynamic balance between tissues and blood vessels. In patients with hypoproteinemia caused by disease or diet, nutritional deficiency in local skin tissues is not conducive to cell growth and wound healing.^18^

In the present study, we found that Lac level was significantly associated with the occurrence of pressure ulcers. Lac is a key product of human glucose metabolism and anaerobic cell metabolism. Increase in Lac level reflects tissue hypoperfusion and anaerobic metabolism. An increase in Lac level and other metabolic disorders caused by ischemia and hypoxia often precede changes in hemodynamics.^19^ Hyperlactatemia is primarily observed in patients with sepsis, patients undergoing cardiopulmonary resuscitation, and those with acute myocardial infarction. Most of these cases are complicated with organ failure and hyperlactatemia. Lactate level is also affected by factors such as shock, ventilator use, and norepinephrine,^20–22^ which are consistent with the disease characteristics and diagnosis of CCU patients. When a patient’s Lac level increases, metabolic abnormalities and microcirculation perfusion often occur and tissue oxygen supply decreases, affecting blood oxygen and nutrition supply to skin tissue.

Vasopressors are commonly used to increase blood pressure and improve perfusion of central organs (brain, heart, lungs, and kidneys) in critically ill patients. However, this class of drugs causes peripheral vasoconstriction and increases the volume of blood returned to the heart while reducing blood flow to peripheral tissues and capillaries. This changes the mechanical characteristics of tissues and heightens their responsiveness to greater mechanical stress (pressure), consequently elevating the risk of pressure injuries.

PCT activates the immune and anti-inflammatory systems of the body during bacterial infection, which leads to the release of endotoxins and an abnormal increase in PCT levels.^23^ PCT can exacerbate inflammatory responses and infections, leading to disease progression. It is also an anti-inflammatory agent involved in the regulation of cytokines.^24^ Some studies^25^ have indicated that PCT is significantly better than C-reactive protein and other inflammatory markers for determining the presence and severity of infection.

Lahmann et al.^26^ showed that diabetes is a risk factor for pressure ulcers. However, diabetes was not identified as a risk factor for HAPIs in the present study. This disparity might have arisen because we employed a binary definition for diabetes (presence or absence of diabetes) and did not gather more detailed diabetes-related information, such as diabetes duration, diabetes type, glycemic control and insulin usage. Another reason is that patients with cardiovascular disease generally have abnormal blood glucose levels, which can be actively and effectively controlled after admission. The correlation between left ventricular ejection fraction and HAPIs has not been reported to date, and no such correlation was observed in the present study.

Clinical variables combined with high-level biomarkers have a superior predictive ability for the occurrence of cardiovascular adverse events. Early risk identification is important^27–28^. Regarding predicting the risk of pressure ulcer in the CCU, we hope to analyze more risk factors in future multicenter studies with large sample sizes to develop a more established and complete prediction model for clinical use.

With the development of artificial intelligence, we hope to develop an intelligent system or method that can automatically extract dynamic data, such as diagnoses, laboratory test results, vital signs, and medical care evaluation records, from the hospital information management system. Automatic dynamic prediction can identify when high-risk patients will likely develop HAPIs to facilitate early initiation of appropriate interventions for them.

## Data Availability

All data produced in the present study are available upon reasonable request to the authors

